# Advancing sarcoma diagnostics with expanded DNA methylation-based classification

**DOI:** 10.1101/2025.06.30.25330543

**Authors:** Natalie Jäger, David E. Reuss, Martin Sill, Daniel Schrimpf, Abigail K. Suwala, Philipp Sievers, Rouzbeh Banan, Felix Hinz, Ramin Rahmanzade, Henry Bogumil, Kaan Fuat Aras, Areeba Patel, Andrey Korshunov, Melanie Bewerunge-Hudler, Arjen HG. Cleven, Manel Esteller, Hanno Glimm, Wolfgang Hartmann, Simon Kreutzfeld, Christoph Heilig, Till Milde, Iver Petersen, Christian M. Vokuhl, Wolfgang Wick, Olaf Witt, Thibault Kervarrec, Evelina Miele, Jonathan Serrano, Stephan Frank, Karl Kashofer, Anne Mc Leer, Elke Pfaff, Melanie Pages, Arnault Tauziede-Espariat, Ferdinand Toberer, Henning B. Boldt, Petr Martinek, Sebastian Brandner, Mayara Euzebio, Aurore Siegfried, Jane Chalker, Patrik Harter, Romain Appay, Wolfgang Dietmaier, Martin Hasselblatt, Uta E. Flucke, Laura S. Hiemcke-Jiwa, David Solomon, Clara Frydrychowicz, Pascale Varlet, Benjamin Goeppert, Michaela Nathrath, Claudia Blattmann, Monika Sparber-Sauer, August Kolb, Michel Mittelbronn, Thomas Mentzel, Sandra Leisz, Anja Harder, Till Acker, Drew Pratt, Eva Wardelmann, Jamal Benhamida, Mark Ladanyi, Philipp Jurmeister, William Foulkes, Pamela Ajuyah, David Z. Ziegler, Jürgen Hench, Maikel JL. Nederkoorn, Yvonne MH. Versleijen-Jonkers, Gunhild Mechtersheimer, Sandro Krieg, Manfred Gessler, Daniel Baumhoer, Sam Behjati, Luca Bertero, Klaus Griwank, Dirk Schadendorf, Pancras CW. Hogendoorn, Jean-Francois Emile, Paul G. Kemps, Armin Jarosch, Michael W. Ronellenfitsch, Toni Su Idler, Daniela Aust, Sylvia Herold, Jessica Pablik, Maysa Al-Hussaini, Zied Abdullaev, Maximus Yeung, Marco Wachtel, Eva Brack, Felix KF. Kommoss, Markku Miettinen, Ken Aldape, Adrienne MH. Flanagan, Uta Dirksen, Kristian Pajtler, Thomas GP. Grünewald, Daniel Lipka, Stefan Fröhling, Christian Koelsche, Matija Snuderl, David Capper, Stefan M. Pfister, David TW. Jones, Felix Sahm, Andreas von Deimling

**Affiliations:** Division of Pediatric Neurooncology, German Cancer Research Center (DKFZ) and German Consortium for Translational Cancer Research (DKTK), Heidelberg, Germany; Hopp Children’s Cancer Center Heidelberg (KiTZ), Heidelberg, Germany; Department of Neuropathology, Institute of Pathology, University Hospital Heidelberg, Heidelberg, Germany; Clinical Cooperation Unit Neuropathology, German Cancer Consortium (DKTK), German Cancer Research Center (DKFZ), Heidelberg, Germany; Department of Pediatric Hematology and Oncology, Heidelberg University Hospital, Heidelberg, Germany; National Center for Tumor Diseases Heidelberg (NCT), Heidelberg, Germany; Genomics and Proteomics Core Facility, German Cancer Research Center (DKFZ), 69120, Heidelberg, Germany; Department of Pathology and Medical Biology, University Medical Center Groningen, Groningen, the Netherlands; Department of Pathology, Amsterdam University Medical Centers, Amsterdam, the Netherlands; Institucio Catalana de Recerca i Estudis Avançats (ICREA), Barcelona, Catalonia, Spain; Physiological Sciences Department, School of Medicine and Health Sciences, University of Barcelona (UB), Barcelona, Catalonia, Spain; Centro de Investigacion Biomedica en Red Cancer (CIBERONC), Madrid, Spain; Cancer Epigenetics Group, Josep Carreras Leukaemia Research Institute (IJC), Badalona, Barcelona, Catalonia, Spain; German Cancer Consortium (DKTK), Dresden, Germany; Translational Functional Cancer Genomics, DKFZ, Heidelberg, Germany; Translational Medical Oncology, National Center for Tumor Diseases Dresden (NCT/UCC), Technical University Dresden and Helmholtz-Zentrum Dresden; Translational Medical Oncology, Technical University Dresden, 01307, Dresden, Germany; Gerhard-Domagk-Institute of Pathology, University of Münster, Münster, Germany; Division of Translational Medical Oncology, German Cancer Research Center (DKFZ), Heidelberg, Germany; Clinical Cooperation Unit Pediatric Oncology, German Cancer Research Center (DKFZ) and German Cancer Consortium (DKTK), Heidelberg, Germany; Department of Pediatric Oncology, Hematology, Immunology and Pulmonology, Heidelberg University Hospital, Heidelberg, Germany; Institute of Pathology, Waldklinikum Gera, Gera, Germany; Department of Pathology, Section Paidopathology, University Hospital Bonn, 53127 Bonn, Germany; Department of Neurology, Heidelberg University Hospital, Heidelberg University, Heidelberg, Germany; Department of Pathology, Centre Hospitalier Universitaire de Tours, Université de Tours, Tours, France; CARADERM Network, Lille, France; Onco-Hematology, Cell Therapy, Gene Therapies and Hemopoietic Transplant, Bambino Gesù Children’s Hospital, IRCCS, Rome, Italy; Department of Pathology, NYU Langone Health and NYU Grossman School of Medicine, New York, New York, USA; Institute for Medical Genetics and Pathology, University Hospital Basel, Basel, Switzerland; Department of Obstetrics & Gynecology and Diagnostic-und Research Institute of Pathology, Medical University of Graz, Austria; Grenoble Alpes University Hospital Pathology Department, Institute for Advanced Biosciences UGA/INSERM U1209/CNRS 5309, Grenoble Alpes University, Grenoble, France; Division of Pediatric Glioma Research, German Cancer Research Center (DKFZ), Heidelberg, Germany; GHU-Paris - Sainte-Anne Hospital, Department of Neuropathology, Paris University, Paris, France; Institut Curie, Paris Sciences Lettres University, SIREDO, INSERM U830, Laboratory of Translational Research in Paediatric Oncology, Paris, France; Department of Neuropathology, GHU Paris - Psychiatry and Neuroscience, Sainte-Anne Hospital, Paris, 75014, France; Department of Dermatology, University of Heidelberg, Heidelberg, Germany; Department of Clinical Research and BRIDGE, University of Southern Denmark, 5000 Odense, Denmark; Department of Pathology, Odense University Hospital, 5000 Odense, Denmark; Bioptical Laboratory, Ltd, Pilsen, Czech Republic; Division of Neuropathology, University College London Hospitals NHS Foundation Trust, Queen Square, London WC1N 3BG, UK; Department of Neurodegenerative Diseases, Queen Square Institute of Neurology, University College London, London WC1N 3BG, UK; Genetics and Molecular Biology, Institute of Biology, State University of Campinas, Campinas 13083-862, SP, Brazil; Research Center, Boldrini Children’s Hospital, Campinas 13083-884, SP, Brazil; Pathology Department, Toulouse University Hospital, Toulouse, France; UFR Santé, Université de Toulouse, Toulouse, France; Toulouse University, Inserm, CNRS, Centre de Recherches en Cancérologie de Toulouse, Toulouse, France; Specialist Integrated Haematology and Malignancy Diagnostic Service-Acquired Genomics, Great Ormond Street Hospital for Children NHS Foundation Trust, London, UK; Institute of Neuropathology, Faculty of Medicine, LMU Munich, Munich, Germany; Neurological Institute (Edinger Institute), University Hospital Frankfurt, Frankfurt am Main, Germany; Department of Pathological Anatomy and Neuropathology, Timone Hospital, APHM, Marseille, France; Institute of Pathology, Center for Molecular Pathology Diagnosis, University of Regensburg, Regensburg, Germany; Institute of Neuropathology, University Hospital Münster, Pottkamp 2, 48149, Münster, Germany; Diagnostic Laboratory, Princess Maxima Center for Pediatric Oncology, Utrecht, The Netherlands; Department of Pathology, Radboud University Medical Center, Nijmegen, The Netherlands; Princess Maxima Center and UMC Utrecht, The Netherlands; Neuropathology, Stanford University School of Medicine, Stanford, USA; Division of Neuropathology, University Hospital Leipzig, 04103, Leipzig, Germany; Institut de Psychiatrie et Neurosciences de Paris (IPNP), UMR S1266, INSERM, IMA-BRAIN, Paris, France; Laboratory of Somatic Genetics, Université de Paris, France3 Curie Institute Hospital, Paris, Paris, France; Institute of Pathology and Neuropathology, RKH Klinikum Ludwigsburg, Ludwigsburg, Germany; Department of Pathology, Heidelberg University Hospital, Heidelberg, Germany; Children’s Cancer Research Centre and Department of Pediatrics, Klinikum rechts der Isar, Technische Universität München, Munich, Germany; Klinikum Stuttgart - Olgahospital, Zentrum für Kinder-, Jugend-und Frauenmedizin, Pädiatrie, Stuttgart, Germany; Klinikum der Landeshauptstadt Stuttgart gKAöR, Olgahospital, Stuttgart Cancer Center, Zentrum für Kinder-, Jugend-und Frauenmedizin, Pädiatrie 5 (Pädiatrische Onkologie, Hämatologie, Immunologie), Stuttgart, Germany; Department of Pathology, New York University Langone Health and School of Medicine, New York, New York, USA; Department of Cancer Research (DoCR), Luxembourg Institute of Health (LIH), 1210 Luxembourg, Luxembourg; Department of Life Sciences and Medicine (DLSM), University of Luxembourg, 4365 Esch-sur-Alzette, Luxembourg; Luxembourg Center of Neuropathology (LCNP), 3555 Dudelange, Luxembourg; Luxembourg Centre for Systems Biomedicine (LCSB), University of Luxembourg (UL), 4362 Esch-sur-Alzette, Luxembourg; Dermatopathology Friedrichshafen, Friedrichshafen, Germany; Department of Neurosurgery, University Medicine Halle, Halle, Germany; Cure NF Research Group, Anatomy and Cell Biology, University Medicine Halle, Halle, Germany; Institute of Neuropathology, Justus-Liebig University Giessen, Giessen, Germany; Laboratory of Pathology, National Cancer Institute, NIH, Bethesda, USA; Department of Pathology and Laboratory Medicine, Memorial Sloan Kettering Cancer Center, New York City, New York, USA; Institute of Pathology, Ludwig Maximilians University Hospital Munich, 80336 Munich, Germany; German Cancer Consortium (DKTK), Munich, Germany; Department of Human Genetics, McGill University, Montreal, Quebec, Canada; Children’s Cancer Institute, Lowy Cancer Research Centre, UNSW, Kensington, NSW, Australia; Kids Cancer Centre, Sydney Children’s Hospital, Randwick, NSW, Australia; Children’s Cancer Institute, Lowy Cancer Centre, UNSW Sydney, Kensington, NSW, Australia; School of Clinical Medicine, UNSW Medicine & Health, UNSW Sydney, Kensington, NSW, Australia; Department of Medical Oncology, Radboud University Medical Center, Nijmegen, The Netherlands; Department of Medical Oncology, Radboud University Medical Centre, 6525 GA Nijmegen, The Netherlands; Department of Neurosurgery, University Hospital Heidelberg, Heidelberg University, 69117 Heidelberg, Germany; Theodor-Boveri-Institute/Biocenter, Developmental Biochemistry, University of Wuerzburg, Wuerzburg, Germany; Comprehensive Cancer Center Mainfranken, University of Wuerzburg, Wuerzburg, Germany; Department of Paediatrics, University of Cambridge; Cambridge, UK; Wellcome Sanger Institute; Hinxton, CB10 1SA, UK; Cambridge University Hospitals NHS Foundation Trust; Cambridge, CB2 0QQ, UK; Pathology Unit, Department of Laboratory Medicine, Department of Medical Sciences, University of Turin, Turin, Italy; Department of Dermatology, University Hospital Essen, University of Duisburg-Essen, Germany & German Cancer Consortium (Deutsches Konsortium für Translationale Krebsforschung, DKTK), Essen, Germany; Comprehensive Cancer Center (Westdeutsches Tumorzentrum), University Hospital Essen, Essen & National Center for Tumor Diseases (NCT) West, Essen, Germany; Research Center One Health, University Duisburg-Essen, Essen, Germany; Department of Pathology, Leiden University Medical Center, Leiden, The Netherlands; Paris-Saclay University, Versailles SQY University, EA4340-BECCOH, Assistance Publique– Hôpitaux de Paris (AP-HP), Ambroise-Paré Hospital, Smart Imaging, Service de Pathologie, Boulogne, France; Department of Pathology, Leiden University Medical Center, Leiden, the Netherlands; Institute of Pathology, Charité - Universitätsmedizin Berlin, Berlin, Germany; Senckenberg Institute of Neurooncology, University Hospital Frankfurt, Frankfurt, Germany; Institute of Pathology, Technical University Dresden, 01307, Dresden, Germany; Department of Pathology and Laboratory Medicine, King Hussein Cancer Center, Amman, Jordan; Department of Pathology, The University of Hong Kong, Queen Mary Hospital, Hong Kong; University Children’s Hospital, Oncology, Balgrist Campus, Zurich, Switzerland; Division of Pediatric Hematology/Oncology, Department of Pediatrics, Inselspital, Bern University Hospital, University of Bern, Bern, Switzerland; The Royal National Orthopaedic Hospital Trust, Brockley Hill, Stanmore HA7 4LP, UK; UCL Cancer Institute, London WC1E 6DD, UK; Pediatric Oncology & Hematology, Pediatrics III, University Hospital of Essen, Essen, Germany; Division of Pediatric Neurooncology, Hopp Children’s Cancer Center Heidelberg (KiTZ); German Cancer Research Center Heidelberg (DKFZ) and Heidelberg University Hospital, Heidelberg; National Center for Tumor Diseases (NCT), Heidelberg, Germany; Division of Translational Pediatric Sarcoma Research, German Cancer Research Center (DKFZ), German Cancer Consortium (DKTK), 69120 Heidelberg, Germany; German Cancer Consortium (DKTK), Heidelberg, Germany; Institute of Human Genetics, Heidelberg University, Heidelberg, Germany; Department of Neuropathology, Charité - Universitätsmedizin Berlin, Berlin, Germany; German Cancer Consortium (DKTK), Berlin, Germany; Pediatric Glioma Research Group, German Cancer Research Center (DKFZ), Heidelberg, Germany

**Author notes:** Contributed equally (Natalie Jäger, David E. Reuss). Correspondence to Andreas von Deimling.

## Abstract

**Purpose:** Sarcomas pose a severe diagnostic challenge. A wide variety of these distinct entities need to be distinguished from each other and from less aggressive types of mesenchymal tumors, to ensure correct clinical management. A machine learning based classifier for sarcomas utilizing DNA methylation data from 1077 tumors recognizing 62 sarcoma types has already been developed and termed the sarcoma classifier, which we published in 2021. Here we present a major advancement of the scale and precision of the sarcoma classifier.

**Methods:** DNA methylation profiles and histologic data from an unprecedented multi-institutional cohort of mesenchymal tumors were collected and analyzed. Utilizing a machine learning approach, the classifier was rigorously validated through five-fold nested cross-validation, achieving a 98% class-level accuracy and a Brier score of 0.017, indicative of well-calibrated probability estimates.

**Results:** The sarcoma classifier v13.1 was developed based on a training set of 4377 methylation profiles from sarcomas and less aggressive mesenchymal tumors comprising 116 tumor sub-classes and 4 control groups forming 93 distinct methylation classes. Performance was validated using four independent cohorts, comprising a total of 1547 mesenchymal tumors. A methylation-based classifier prediction was obtained in 73% of cases in the validation sets, of which 91% matched the original histopathology diagnosis, thereby increasing diagnostic confidence. The classifier enabled a definitive molecular diagnosis or tumor reclassification in 6% of cases with inconclusive or ambiguous histological findings.

**Conclusion:** Adding new sarcoma types and expanding tumor sample numbers in each methylation class in the new sarcoma classifier decisively increased the number of diagnostic predictions and improved match with histologic evaluation. This substantial advancement will promote clinical implementation of the tool for the diagnosis of mesenchymal tumor lesions.

## Introduction

The challenges of diagnosing sarcomas result in a high interobserver variation, requiring of reevaluation by referral centers, and the performing of an increasing array of different molecular analyses ^1,2^. Typical molecular analyses focus on DNA and RNA sequencing or FISH in order to find specific genetic alterations. Approximately half of the sarcoma types exhibit highly characteristic gene fusions, and therefore, cDNA sequencing is of utmost relevance in these tumors. Another molecular approach to sarcoma classification has previously been established using DNA methylation data, leveraging insights gained from the brain tumor classifier published in 2018 ^3^. Subsequently, a sarcoma classifier version v12.3 had been developed, which recognized 62 tumor methylation classes covering already a wide spectrum of mesenchymal tumors ^4^. In contrast to sequencing approaches searching for pathognomonic mutations, the methylation-based approach aims at identifying methylation patterns which are closely related to the cell of origin of the tumor cells. This implies the need for a comprehensive sample reference set able to cope with the intertumoral variance of DNA methylation patterns. Expanding the reference dataset allows for broader representation of the biological diversity within and across tumor entities, which may facilitate the identification of additional methylation classes and increase the confidence and specificity of classifier predictions. To this end we have advanced the existing tool to the new sarcoma classifier v13.1 by adding 62 additional tumor types or subtypes. In addition, our hierarchical system leverages the well-calibrated classifier probability scores by aggregating subclass-level scores at higher tiers, thereby providing more robust diagnostic guidance at higher-order levels. A total of 120 subclasses is covered in 89 different tumor methylation classes and 4 control classes, based on a reference set containing 4377 DNA methylation array profiles (Fig. 1).

**Figure 1.**
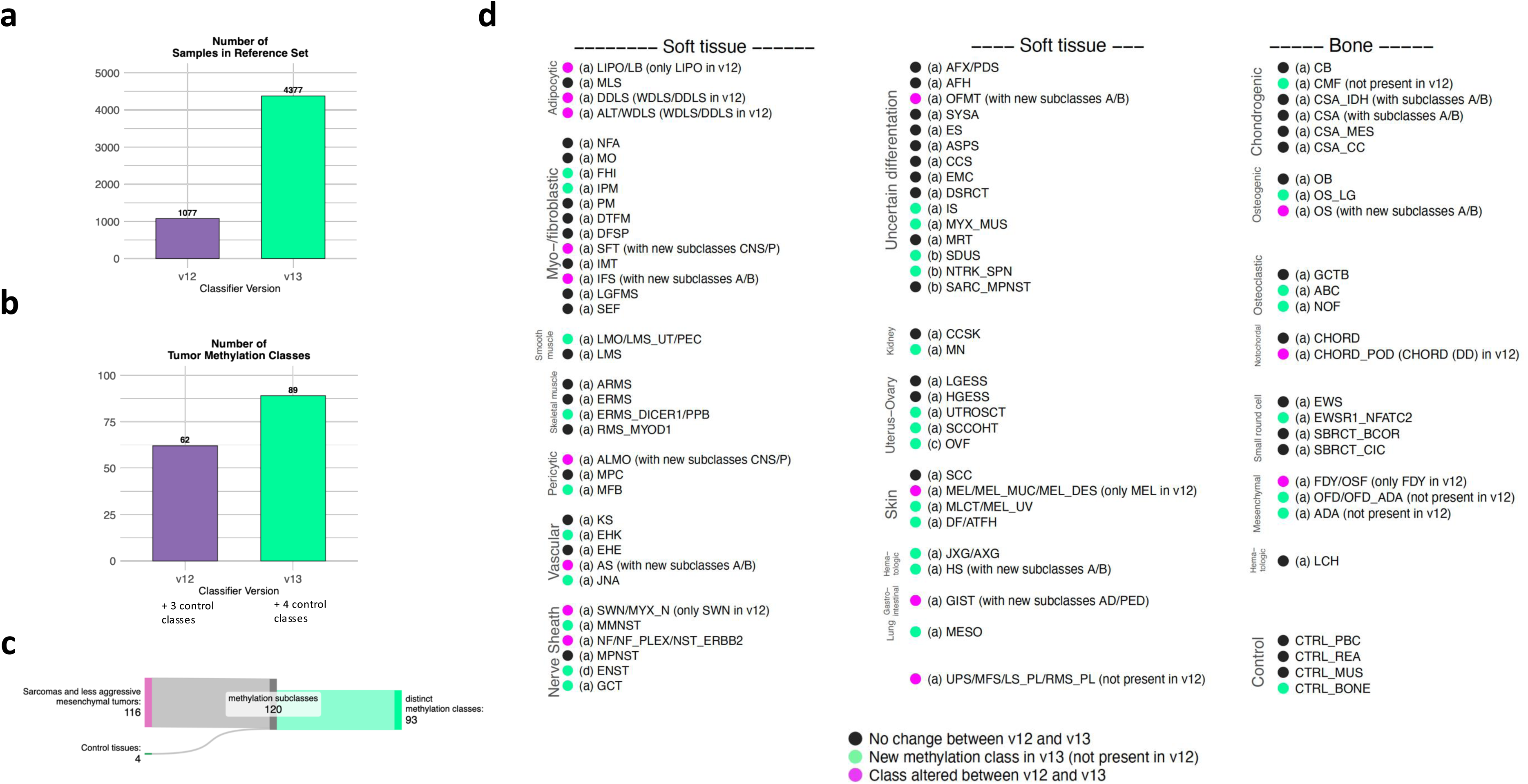
Comparison of the sarcoma classifier versions v12.3 and v13.1. (a) The size of the reference dataset was expanded from 1077 to 4377 samples. (b) The number of tumor methylation classes increased from 62 in v12.3 to 89 in v13.1, including additional control classes. (c) Sankey diagram to illustrate how the reference dataset from sarcomas and less aggressive mesenchymal tumors comprising 116 tumor and 4 control groups forms 93 distinct methylation classes. (d) Overview of tumor types and subtypes included in v13.1, highlighting refined subclass definitions, additions, and restructuring compared to v12.3. Notably, several entities were subclassified (e.g., AS, SFT, OFMT, GIST), while others were newly introduced (e.g., CMF, ABC, ADA).

### Material and Methods

#### DNA-Methylation Array Processing

The Illumina Infinium HumanMethylation450 (450k) array, Illumina Infinium MethylationEPIC (EPIC) array and Illumina Infinium MethylationEPICv2 (EPICv2) microarrays were used to obtain genome-wide DNA methylation data for tumor samples and normal control tissues according to the manufacturer’s instructions (Illumina, San Diego, USA). DNA methylation data were generated from both fresh-frozen and formalin-fixed paraffin-embedded (FFPE) tissue samples. For most of the fresh-frozen samples and FFPE tissue samples, between 50 ng and 500 ng of DNA was used as input material. On-chip quality metrics of all samples were carefully controlled.

All computational analyses were performed in R version 4.3.3 (R Development Core Team, 2024). Raw signal intensities were obtained from IDAT files using the minfi Bioconductor package version 1.21.4 ^5^. Illumina EPIC, EPICv2 and 450k samples were merged into a combined dataset by selecting the intersection of probes present on all arrays (combineArrays function in minfi). Each sample was individually normalized by performing a background correction (shifting of the 5% percentile of negative control probe intensities to 0) and a dye-bias correction (scaling of the mean of normalization control probe intensities to 10,000) for both color channels. Subsequently, a correction for the type of material (FFPE/frozen) and array type (450k/EPIC(v2)) was performed by fitting univariable linear models to the log2-transformed intensity values (removeBatchEffect function in the limma package version 3.30.11). The methylated and unmethylated signals were corrected individually. Beta-values were calculated from the retransformed intensities using an offset of 100, as recommended by Illumina.

#### Methylation Data Quality Control

The so-called “detectionP” is based on detection p-values for all probed genomic positions (https://rdrr.io/bioc/minfi/man/detectionP.html). The detection p-value threshold indicates the probability that the observed signal is due to background noise. A cutoff of p < 0.05 is usually recommended for detection p-values. *Minfi* further provides a quality control metric that uses the log median intensity in both the methylated (M; Median Log2 Meth) and unmethylated (U; Median Log2 Unmeth) channels. When plotting these two medians against each other, good samples tend to cluster together, while failed samples tend to separate and have lower median intensities.

The reference training data set as well as the validation data sets were filtered to contain only samples, which exceed the following three data quality-relevant control metrics: “Fraction of detectionP < 0.05” > 0.95; Median Log2 Meth > 8; Median Log2 Unmeth > 8. Further, all samples of the data set indicating matching genotypes based on the SNPs on the methylation arrays were also de-duplicated to prevent having included the same patient sample more than once.

#### Classifier training

Classifier training was performed as described previously ^3,6^ . First, we applied a permutation-based variable importance measure (R-package randomForest v4.7-1.2) to select the 10,000 most informative CpG probes as features for the final Random Forest (RF). Unbalanced class sample sizes were considered by down sampling each bootstrap sample to the minority class. Next, a ridge-penalized multinomial logistic-regression model (R-package glmnet v4.1-8) was fitted to calibrate the RF output, mapping raw prediction scores to probability estimates. An optimal penalization parameter was chosen by a ten-fold cross-validation. Combining classifier outputs with a logistic regression model is an ensemble strategy known as stacking^51^.

#### Classifier validation

To evaluate the classifier, a five-fold nested cross-validation scheme generated out-of-sample RF scores that enabled us to fit and validate the calibration models in each fold. To measure the performance of the classifier the following metrics and figures were generated: Accuracy, Balanced Accuracy, F1, Matthews Correlation Coefficient (R-package mltest v1.0.1), Confusion Matrix, multiclass Log Loss, multiclass Brier Score, and Calibration Plots (R-package rms v6.8-2). In addition, receiver operating characteristics (ROC) curves and accompanying areas under the curve (AUC) were generated by binarizing multiclass predictions (correct vs. incorrect) (R-package pROC v1.18.5).

#### Estimating tumor cell content from DNA methylation data

Tumor cell content for all reference and validation samples was estimated using a deconvolution approach based on a previously published reference signature matrix developed for immune cell deconvolution in CNS tumors (https://pubmed.ncbi.nlm.nih.gov/32859926). This matrix was derived from DNA methylation profiles of pure, flow-sorted cell populations, including profiles from cancer cell lines representative of CNS tumors and malignant rhabdoid tumors (MRT).

In addition, for samples analysed with the HumanMethylation450 (450k) or Illumina Infinium MethylationEPIC (EPIC) array, the R package RF_Purify was used to estimate the tumor cell content (EPICv2 samples cannot be analysed with RF_Purify). The tumor cell content estimates from both methods were well correlated (Suppl. Fig 4).

#### CNV analysis

Gene amplifications and homozygous deletions (deep deletions) were called according to a published algorithm (https://forum.depmap.org/t/defining-deep-deletions-and-amplifications/710/5). Values lower than -1.2 on a log scale were scored as homozygous deletions and values higher than 0.75 on a log scale were scored as amplification.

## Results

### Compiling the sarcoma classifier reference set

The present dataset substantially expands upon the reference set of the established sarcoma classifier^4^. While v12.3 was generated on cases with morphological expert review and extensive molecular workup, the updated classifier version employs a combined approach, incorporating both molecularly well-annotated cases and cases selected based on matching diagnoses aligned with their association to specific methylation-defined clusters. The number of reference cases including controls increased from 1077 to 4377 cases (Fig. 1a), which have all been subjected to t-Distributed Stochastic Neighbor Embedding (t-SNE) and to Uniform Manifold Approximation and Projection (UMAP) in order to ensure their association with distinct methylation classes. Of the 62 tumor methylation classes from the sarcoma classifier v12.3 (Fig. 1b), 47 remained unchanged while 15 were modified either by subdivision or by combining with additional tumor methylation classes. The updated sarcoma reference set v13.1 now includes 55 new tumor classes not represented in the initial version (Fig. 1d). Sample numbers in distinct methylation classes ranged from 6 to 205 cases. Full descriptions matching to the abbreviations of methylation classes and information on case numbers within each methylation class is provided in supplementary table 1. Controls included, the updated reference set contains 93 methylation classes relating to 124 tumor and four control groups (Figure 1b, c). The 128 tissue types contain 117 mesenchymal tumors, four controls and non-mesenchymal tumors including SCC (squamous cell carcinoma (cutaneous)), LCH (Langerhans cell histiocytosis), HS (histiocytic sarcoma), MEL (melanoma), MEL_MUC (melanoma mucosal) and MEL_DES (melanoma desmoplastic). The non-mesenchymal tumors were included, because they constitute relevant differential diagnoses to several sarcoma types. One entity, the SCCOHT (small cell carcinoma of the ovary, hypercalcemic type), was initially considered an epithelial tumor but more recently is discussed in the context of sarcomas or primitive germ cell tumors ^7^. Brain specific sarcomas such as gliosarcoma, oligosarcoma, and primary intracranial sarcoma DICER1-mutant were not included, because they are represented in the organ specific brain tumor classifier v12.8 (https://app.epignostix.com/#/classifiers). Further including these sarcomas would have required to also include oligodendrogliomas, glioblastomas and other brain tumors into the sarcoma classifier v13.1 for clear separation of these tumor types. The methylation classes are briefly described in supplementary table 2.

All 4377 tissue samples constituting the reference set have been analyzed for chromosomal alterations manifesting as gains and losses and for amplifications and homozygous deletions in a specific gene set. An overview of the CNV distribution in all tumor methylation classes is given in supplementary Fig. 2. To provide guidance on the available information on the new and existing classes and subclasses and their alignment with the current WHO classification, we introduced an evidence level annotation as specified in Fig.1d and Fig. 2a and supplementary table 2: Level a - Tumor type/subtype identical to WHO 2020; Level b - Large single or more than one smaller dataset published describing the type/subtype as molecularly and/or clinically distinct, or the methylation class represents a distinct fraction of an established WHO 2020 tumor class; Level c - Single small dataset or case series; Level d- solely based on clusters in tSNE/UMAP. The annotation is typically provided at the most granular layer, plus in part also at a higher layer if that instead matches a WHO type/subtype.

**Figure 2.**
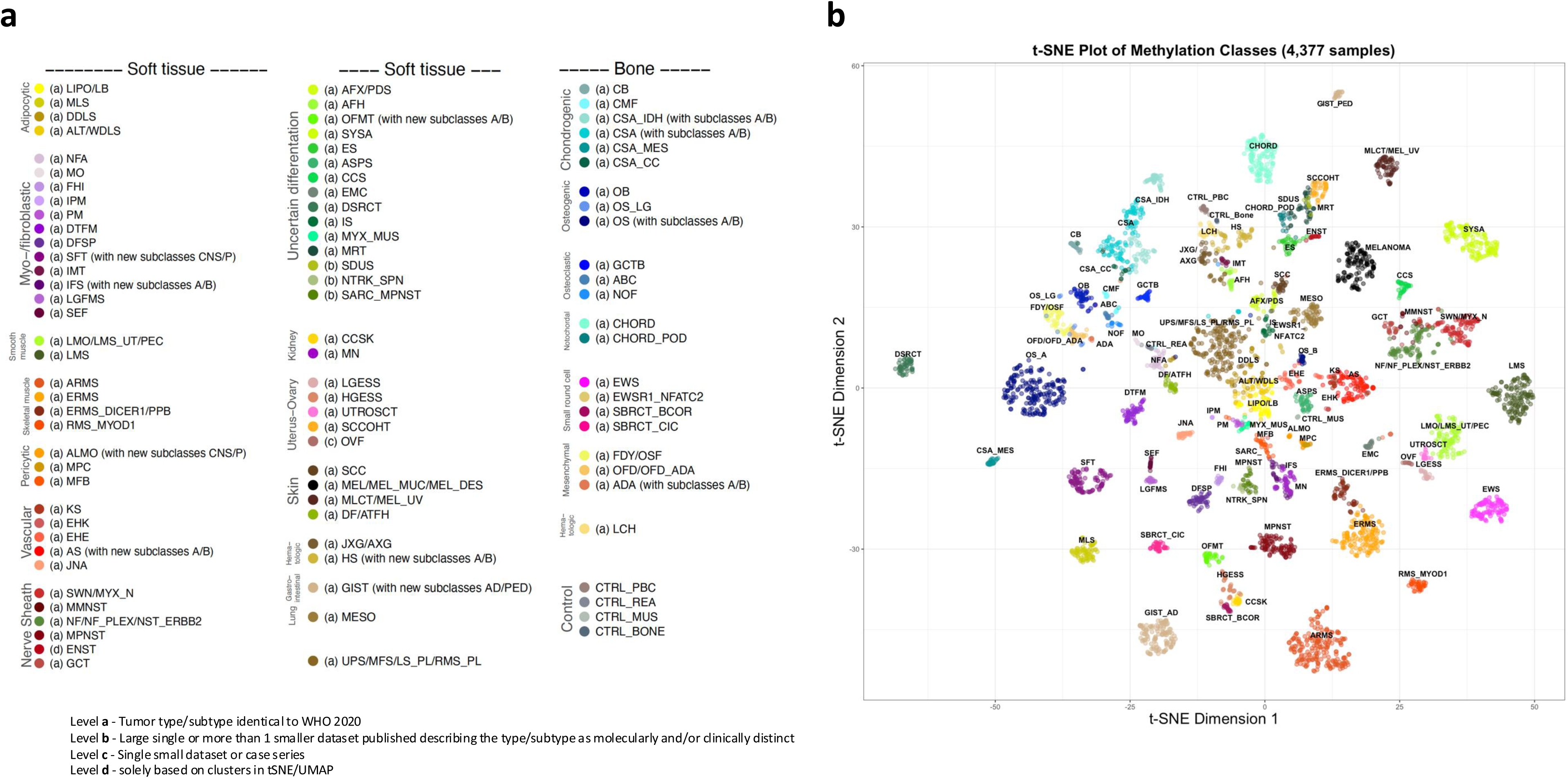
Overview of the DNA methylation-based sarcoma reference cohort used for training the v13.1 sarcoma classifier. (a) Overview of 89 tumour and four control DNA methylation classes included in the v13.1 sarcoma classifier reference cohort. The sarcoma methylation classes are grouped according to the WHO 2020 scheme. Letters in rounded brackets before abbreviation of the class indicate ‘evidence level’ for the respective class. Full names and further details of the methylation classes are given in supplementary tables 1 and 2. (b) t-SNE projection of complete reference sample set (4377 DNA methylation profiles) used for training the v13.1 sarcoma classifier.

### Classifier v13.1 generation and cross validation performance

Unsupervised clustering using t-SNE and UMAP, along with cross-validation results, revealed substantial overlap in the methylation profiles of a small number of tumor types and subtypes. Due to these overlapping methylation profiles, reliable separation of certain entities was not feasible. Therefore, seven combined methylation classes were defined by grouping these entities for classifier training, even though they do not represent established tumor subclasses. These combined methylation classes included only non-sarcoma tumors that are relevant in the differential diagnosis of sarcomas, such as melanomas (supplementary table 1). In summary, the v13.1 sarcoma classifier was trained using 116 subclasses and four groups for normal control tissue (Fig. 1c).

The updated sarcoma classifier model was developed using a Random Forest–based approach ^8^ as previously described ^3,4^. To evaluate its performance, the classifier was validated using a five-fold nested cross-validation scheme. All subclasses achieved a balanced accuracy greater than 0.64, with 101 out of 120 subclasses exceeding 0.9 in accuracy (Fig. 3a). The newly introduced hierarchical system leverages these well-calibrated probabilities by aggregating subclass-level scores at higher tiers, specifically for the subclasses with lower accuracy, thereby providing more robust diagnostic guidance at these higher-order tiers. Most of the more granular subclasses currently lack direct clinical implications, making aggregated probability scores at the higher Class level sufficient for guiding diagnostic decisions.

**Figure 3.**
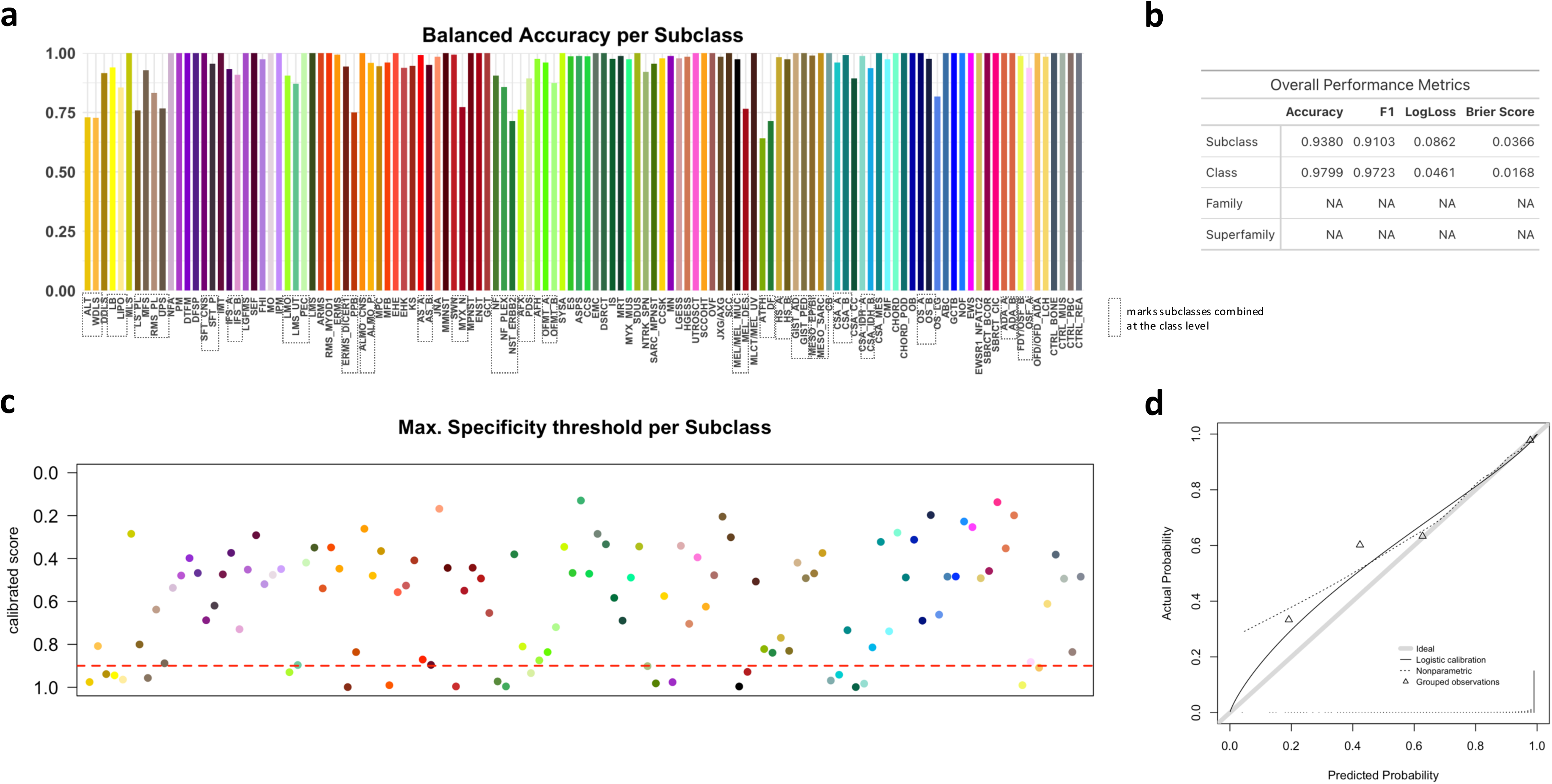
Performance of the v13.1 sarcoma classifier in five-fold nested cross-validation. (a) Bar plot showing the balanced accuracy for each of the 93 subclasses, as derived from five-fold nested cross-validation. Subclass combinations into classes are indicated where applicable by a box with dashed lines. (b) Table summarizing overall performance metrics—accuracy, F1-score, log loss, and Brier score—evaluated at each hierarchical level (subclass, class). (c) Scatterplot of subclass- specific maximum specificity thresholds with color-coded subclasses and a red dashed line at 0.9 marking the recommended threshold for the calibrated score. (d) Calibration plot comparing predicted probabilities with observed outcomes, illustrating the degree of score calibration across subclasses.

Overall, the classifier achieved a 98% Class-level accuracy with a Brier Score of 0.0168, indicating well-calibrated probability estimates (Figure 3b).

As with the previous v12.3 version, we recommend a score threshold of 0.9 at the Class level as a “high-confidence prediction” for the v13.1 sarcoma classifier (Fig. 3c, d). This threshold offers a practical balance between clarity in communication and robust performance in clinical settings. To identify statistically optimal thresholds, we performed ROC analyses for each subclass in a one-against-all framework and computed, for each subclass, the threshold that maximizes the specificity (Fig. 3c). While the optimal thresholds ranged from as low as 0.13 up to 0.99—indicating that certain classes, such as adipocytic sarcomas, may require higher cutoffs for maximal specificity—the 0.9 threshold still ensures high specificity for the vast majority of classes (max. specificity mean of all subclasses = 0.6; Fig. 3c).

### Methylation classes comprising several tumor entities

The v13.1 reference set did not allow for clear separation of all tumor subclasses included, resulting in several methylation classes containing two or more tumor entities. Many of these methylation classes combined tumors assumed to be highly related to each other. Examples are methylation classes (mc) AFX/PDS (atypical fibroxanthoma/ pleomorphic dermal sarcoma) ^9^, mc DF/ATFH (dermatofibroma/ (atypical fibrous histiocytoma), mc JXG/AXG (juvenile/adult xanthogranuloma), mc MESO (mesothelioma) and mc ALT/WDLS (atypical lipomatous tumor/ well differentiated liposarcoma), the latter separated on the basis of tumor location and potential respectability ^10^ with both tumor types having a high incidence of *MDM2* amplifications. Several methylation classes contain lesions of different morphology or tumor localization but sharing similar driver mutations such as mc MLCT/MEL_UV (melanocytoma/ uveal melanoma) with *GNAQ* or *GNA11* mutations, and mc ERMS_DICER1/PPB (embryonal rhabdomyosarcoma DICER1-mutant / pleuropulmonary blastoma) with *DICER1* mutations.

Common to tumors in mc NF/NF_PLEX/NST_ERBB2 (neurofibroma / neurofibroma plexiform / nerve sheath tumour, ERBB2-mutant) is growth in peripheral nerves and *NF1* mutations in NF and NF_PLEX while NST_ERBB2 are characterized by mutations in the receptor tyrosine kinase gene *ERBB2*. The mc MEL/MEL_MUC/MEL_DES (melanoma / melanoma-mucosal/melanoma demoplastic) comprises cutaneous, mucosal and desmoplastic melanoma. The mc FDY/OSF (fibrous dysplasia/ ossifying fibroma) combines two lower grade osseous lesions. Of these lesions, FDY typically carry *GNAS* mutations. The mc LMO/LMS_UT/PEC (leiomyoma / leiomyosarcoma (uterine type) / PEComa) combines tumors of seemingly different morphology and grades, separating the uterine leiomyosarcomas from their counterparts in other localizations. Several sarcomas subsumed under the term “pleomorphic sarcomas” ^11^ comprising UPS, MFS, pleomorphic liposarcoma and pleomorphic rhabdomyosarcoma are combined in mc UPS/MFS/LS_PL/RMS_PL. RMS_PL (undifferentiated pleomorphic sarcoma / myxofibrosarcoma / pleomorphic liposarcoma / pleomorphic rhabdomyosarcoma) and UPS as well as LS_PL and MFS have been shown to exhibit similar genomic alterations ^12,13^ which is supported by a similar CNV profile of these sarcomas (supplementary Fig. 2). The mc OFD/OFD_ADA (osteofibrous dysplasia / osteofibrous dysplasia like adamantinoma) is clearly separate from mc ADA supporting previous findings on molecular differences between osteofibrous dysplasia-like adamantinoma and adamantinoma ^14^.

### New subclasses within sarcoma entities based on DNA methylation

Angiosarcoma (AS) was split up into two methylation subclasses. The subclass AS_A was characterized by a high frequency of *MYC* amplifications (54%), which were not observed in subclass AS_B (Suppl. Fig.1a). Similar to a previous report, *MYC* amplification occurred in AS arising in different localizations^15^. Radiation associated AS accumulated in subclass AS_A. No specific clinical feature was associated with AS_B. The number and extent of chromosomal alterations was larger in AS_A.

Two separate methylation subclasses for gastrointestinal stromal tumour (GIST) were detected in the reference set, termed GIST_AD (median age 64 years, AD=adult type) and GIST_PED (median age 15 years, PED=pediatric type). The mc GIST_PED likely corresponds to the (*KIT or PDGFRA*) wild-type GIST preferentially seen in young patients supported by the lack of chromosome 14 deletions and a high rate of *MGMT* gene methylation in this group ^16^ and a marked female predominance (Suppl. Fig.1b). Among osteosarcoma (OS), a small group of 21 cases termed OS_B separated from the bulk of 189 cases termed OS_A. Copy number variations were clearly lower in OS_B than in OS_A, as well as compared to low grade central osteosarcoma (supplementary Fig. 2). Currently, no clinical outcome data are available for osteosarcoma class OS_B.

Patients with histiocytic sarcoma (HS) belonging to mc HS_A and HS_B did not differ in regard to age and tumor localization, however CNV alterations were more numerous in HS_A than in HS_B. Infantile fibrosarcoma (IFS) presented with two distinct methylation classes, IFS_A and IFS_B. Of these, IFS_A formed a cluster together with mesoblastic nephroma (MN) while IFS_B was separate. Both IFS subgroups exhibited comparable CNVs with gains on chromosomal arms 11p/q 15,17p/q and 19p/q being most frequent. Interestingly, gains on chromosome 11 were also shared with MN of the cellular subtype, which show recurrent *ETV6-NTRK3* fusions.

MPNST included a histologically recognized variant frequently diagnosed as epithelioid MPNST, which exhibits a characteristic methylation profile and typically carries chromosome 2 gains or *SMARCB1* homozygous deletions, suggesting the presence of a distinct tumor type provisionally termed epithelioid nerve sheath tumor (ENST). This group shows overlap with a previously reported cohort ^17^, although ENST do not exhibit malignancy in all cases.

OFMT (ossifying fibromyxoid tumour) displays two different methylation profiles for subclasses OFMT_A and OFMT_B. No distinct clinical characteristics for these methylation subgroups have been defined yet. However, the different CNV profiles for OFMT_A and OFMT_B (supplementary Fig. 2) point towards differences beyond only the methylation profile.

### Sarcoma subgroups depending on tumor localization

Four tumor types, that is angioleiomyoma (ALMO), solitary fibrous tumor (SFT), schwannoma (SWN) and IDH-mutant chondrosarcoma exhibited anatomic location-specific methylation patterns with intracranial tumors clustering separately from peripheral tumors (Fig. 2b). Site specific clustering was also observed in leiomyosarcoma, with uterine tumors clustering differently from those in other localizations. In addition to the instances described here, separation of methylation classes based on tumor localization was also previously observed for several brain tumor entities, such as pilocytic astrocytoma and subependymoma. These tumors tended to cluster depending on respective brain compartments ^3^. This indicates the importance of tumor location and possible microenvironmental effects and may be of relevance for future classifiers.

### MDM2 amplification status and DDLS prediction

All liposarcomas in the reference set of v13.1 exhibited *MDM2* amplifications. We observed classifier prediction of UPS/MFS/LS_PL/RMS_PL in some DDLS cases exhibiting *MDM2* amplification. Other tumors with *MDM2* amplification were not affected, since their respective methylation profiles were sufficiently different from DDLS. Therefore, we included a caveat in the classification output of v13.1 pointing towards the possibility of DDLS if CNV analysis recognized *MDM2* amplification. Specifically, to further support a DDLS classifier prediction, we recommend reviewing the Copy Number Variation (CNV) profile of the tumor for evidence of *MDM2* gene amplification.

### Independent validation of the v13.1 classifier in external sarcoma cohorts

Validation of the v13.1 sarcoma classifier was performed employing four independent tumor data sets with a total of 1547 cases. Prior to analyses, all data sets had to pass stringent quality checks. Cases with quality-relevant values below our described thresholds were excluded from the analysis, resulting in a total of 1547 cases passing all quality metrics (see Materials and Methods for details). Samples were defined as within classifier scope (predictable or eligible) if their original histopathological diagnosis aligned with one of the tumor (sub-)classes included in the v13.1 reference cohort.

High tumor cell content positively correlated with more cases reaching a classifier prediction (supplementary Fig. 3a). In contrast, the likelihood for a confident classifier prediction to match the original histopathological diagnosis is less dependent on tumor cell content (supplementary Fig. 3b). All cases from the four separate validation data sets are listed in supplementary table 3. A graphic overview, including the distribution of original diagnoses of the validation cohort, is provided in Fig 4a and b.

**Figure 4.**
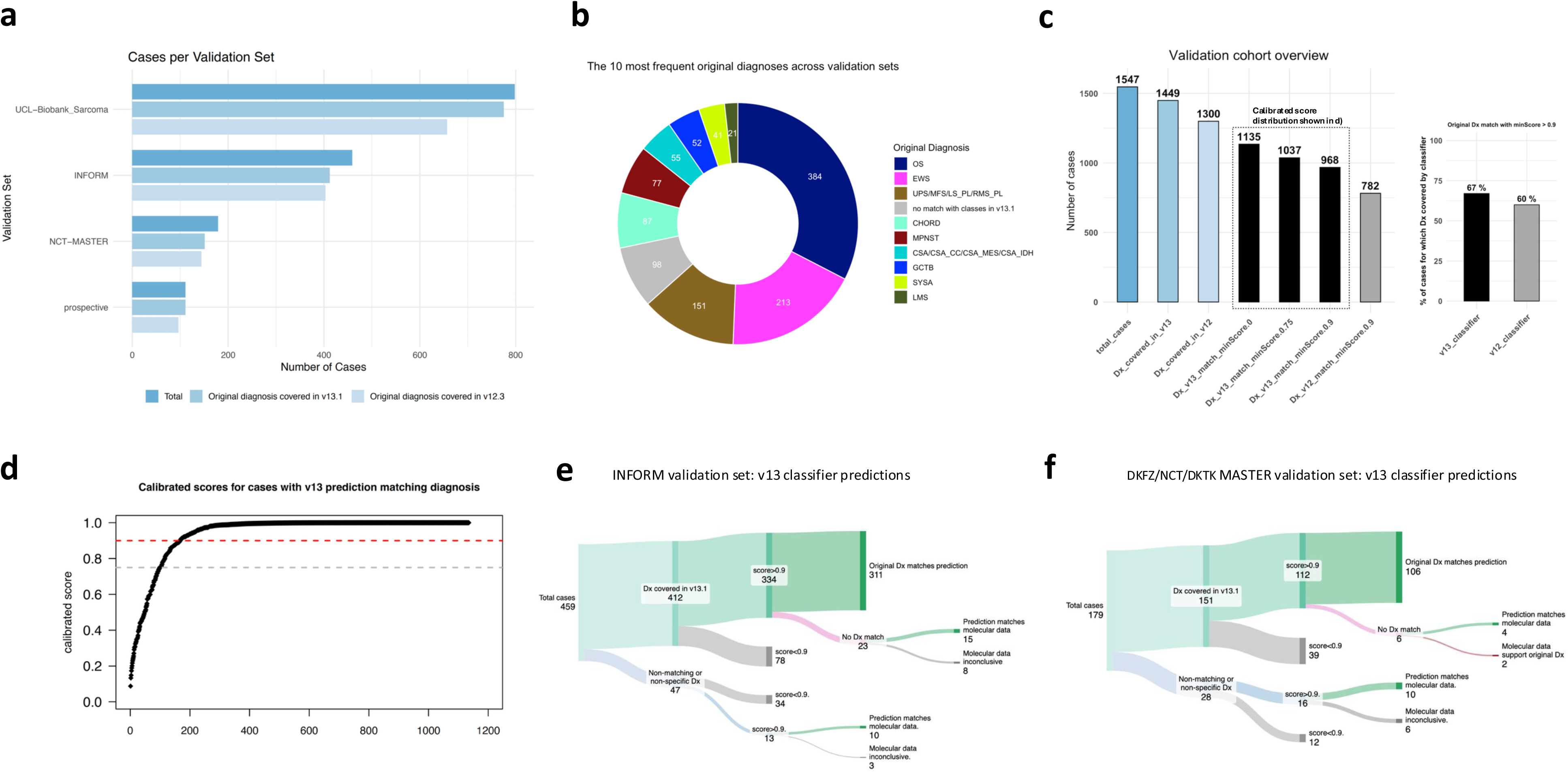
Independent validation of the v13.1 classifier in external sarcoma cohorts. The classifier was evaluated on 1547 independent samples from four different sources: DKFZ/NCT/DKTK MASTER, INFORM, UCL Biobank, and a prospective cohort from TU-Dresden university hospital. (a) Distribution of cases by source cohort. (b) The 10 most frequent original (institutional) diagnoses across the combined validation cohort. Numbers shown within the donut chart represent the number of samples per diagnosis. (c) Overview of v13.1 sarcoma classifier predictions matching the original diagnoses with different calibrated score thresholds (black bars) across the combined validation cohort, and comparison with the previous sarcoma classifier version (v12.3, grey bars) (d) Distribution of calibrated classifier scores for cases with v13.1 classifier prediction matching original diagnosis, and a red dashed line at 0.9 marking the recommended score threshold. (e) Sankey diagram showing concordance between v13.1 classifier predictions and original diagnoses in the INFORM validation data set, considering the molecular data (NGS-based data for each tumor) for discrepant cases. (f) Sankey diagram showing concordance between v13.1 classifier predictions and original diagnoses in the DKFZ/NCT/DKTK MASTER validation data set, considering the molecular data (NGS- based data for each tumor) for discrepant cases, resulting in classifier-based refinement of original diagnoses for 14 cases in total. Dx = Diagnosis.

All 1547 cases from the four validation cohorts were pooled, yielding 1300 cases with tumor types covered by the methylation classes of classifier version v12.3, and 1449 such cases for version v13.1 (Fig. 4c). For all cases of the validation data set for which the v13.1 prediction matches the original diagnosis, the calibrated scores for these predictions exceed the recommended threshold of 0.9 in 968/1135 (85%) cases (Fig. 4c, d).

With the v12.3 classifier, 864 predictions and with the v13.1 classifier, 1061 predictions were reached corresponding to 66% and 73% predictions in cases, for which original histopathological diagnosis aligned with one of the tumor classes included in the v13.1 reference cohort. Concordant diagnosis in eligible cases with predictions (score > 0.9) were obtained in 90,5% employing v12.3 and 91,2% employing the new v13.1 classifier version.

#### INFORM cohort

The INFORM cohort enrolls pediatric patients with predominantly recurrent tumors for extensive molecular testing including DNA sequencing, RNA sequencing and methylation analysis ^18^. 459 cases of the INFORM cohort with a sarcoma diagnosis were included in the validation set. Of these, 47 cases did not match in their histopathological diagnosis with the tumor types represented in the v13.1 reference set, or the initial diagnosis was non-specific, like “Soft Tissue Sarcoma” for example. INFORM sarcoma samples already included in the reference set were not included in the validation set.

Being restricted to pediatric patients, the most frequently represented sarcoma type was Ewing sarcoma followed by osteosarcoma. Data on individual cases were compiled in supplementary table 3. In 311 out of 459 cases (68%), the histopathological diagnosis was supported by a classifier prediction with a score greater than 0.9, thereby increasing diagnostic confidence (Fig. 4e). The classifier enabled a definitive molecular diagnosis or tumor reclassification in 25 of 459 (5%) cases with inconclusive or ambiguous histological findings (Fig. 4e), providing added value on top of current standard diagnostic practice.

#### MASTER cohort

The prospective DKFZ/NCT/DKTK MASTER trial (ClinicalTrials.gov: NCT05852522) enrolls adult patients with advanced rare cancers who have exhausted standard treatment options to undergo extensive molecular testing, including whole-genome/exome sequencing, transcriptome sequencing, and DNA methylation profiling, to inform clinical management. ^19^. Of the 179 cases in total, 151 were considered eligible (histopathological diagnosis covered in the v13.1 classifier) and among these 75% received a high-score prediction. In 94,6% of the cases, the prediction matched the original diagnosis (Fig. 4f). Reevaluation of the clinical diagnosis for 6 non-matching patients based on molecular findings resulted in a change of diagnosis in four patients (marked * in supplementary table 3). Two discrepancies concerning issues in liposarcoma prediction by v13.1 could be resolved based on *MDM2* amplification in both cases. Thus, the sarcoma classifier version v13.1 predicted 98,0% of cases correctly in a validation set with strict molecular workup of initially discrepant cases. In the two mismatching cases, v13.1 explicitly hinted towards the problem and suggested consideration of the correct diagnosis of liposarcoma (DDLS). In addition, ten cases for which their original histopathological diagnosis was not included in the v13.1 reference cohort received an updated diagnosis based on molecular data. The updated diagnosis and the v13.1 classifier prediction matched in these 10 cases. In summary, in 106 out of 179 cases (60%), the histopathological diagnosis was supported by a classifier prediction with a score greater than 0.9, thereby increasing diagnostic confidence (Fig. 4f). The classifier enabled a definitive molecular diagnosis or tumor reclassification in 14 of 179 (8%) cases with inconclusive or ambiguous histological findings (Fig. 4f).

#### UCL Biobank sarcoma set

This set is based on publicly accessible data which we have utilized to validate the first version (v12.3) of the sarcoma classifier ^20^. The series predominantly contains sarcomas involving bone and cartilage with osteosarcoma and chondrosarcoma being most frequently represented. From the series of 798 cases in total, 775 cases were eligible with a prediction reached in 69%. Prediction and diagnosis matched in 88.9 % of the cases. An overview is given in suppl. table 3.

#### Prospective cohort

A series of 111 cases was compiled from prospective new diagnoses after finalizing the v13.1 sarcoma classifier. Only cases with their initial diagnoses represented in the v13.1 sarcoma classifier classes were included. A prediction (with classifier score > 0.9) was reached in 74 % of the cases with 93.9 % of the predictions concordant with the initial diagnosis. An overview is given in supplementary table 3.

## Discussion

Methylation based tumor classification has proven highly valuable in human brain tumors ^3^. Non-neuroectodermal tumors, however, may pose a greater challenge for applying this approach than brain tumors. One emerging explanation is the previously underestimated influence of tissue-specific factors on the methylation pattern, as exemplified by the growing number of distinct methylation classes observed for a single tumor entity arising in different organ systems. Typical examples are SFT (solitary fibrous tumor), ALMO (angioleiomyoma) and schwannoma (SWN) with distinct methylation patterns depending on their tumor origin in- or outside the CNS. Two hypotheses may suggest potential origins for the localization specificity observed in methylation classes within tumor entities: firstly, the tumor precursor cells from distinct anatomic locations may exhibit sufficient differences in their methylation pattern to be recognized by the classifier algorithm. Second, site-specific portions of non-tumorous tissues, i.e. the microenvironment, contribute significantly to the methylation pattern. This may require a site-specific approach, with organ-specific classifiers for the vast majority of epithelial tumors, but possibly also for sarcomas, constituting the best solution for future algorithms. In fact, future organ-specific classifiers may even combine epithelial and mesenchymal tumors.

Version v12.3 ^4^ of the sarcoma classifier has been subject to multiple independent testing ^20–22^. While in general considered to be a very promising diagnostic aid, a common concern was the high rate of unclassifiable cases. To this end, v13.1 was expanded to include 93 methylation classes in total, adding 62 new tumor types or subtypes to the previous version. In four validation sets totaling 1547 cases, the rate of predictions increased from 66% in v12.3 to 73% in v13.1. The rate of concordance of prediction and pathological diagnosis increased slightly from 90,5% to 91,2%. The v13.1 classifier comprises more methylation classes that include several tumor types or subtypes at the more fine-grained methylation subclass level than the predecessor, the v12.3 sarcoma classifier. Some entities such as myxofibrosarcoma or desmoplastic melanoma did not separate well from undifferentiated pleomorphic sarcoma or melanoma, respectively, and therefore were not included in v12.3. Inclusion of these entities in v13.1 necessitated the formation of hierarchical methylation classes with contribution from more than a single tumor group. It was thus considered that predicting combined methylation classes based on one or more subordinate subclasses would reduce the risk of misclassification, with further distinction remaining in the expert hands of the pathologist. This approach underlines the concept that the sarcoma classifier is a tool for suggesting diagnostic directions and for aiding decision making, rather than providing a concrete final diagnosis.

The new sarcoma classifier was validated using 1527 tumor samples from four independent sarcoma sets of different composition with respect to tumor types. The rate of diagnostic predictions rose to 73% in v13.1 from 66% in v12.3. Of relevance is a strongly increased rate of failed predictions in tumor samples with less than 40% tumor cell content, pointing to the problem of sampling best suited tumor material prior to downstream molecular analysis. The rate of matched predictions increased slightly from 90,5% in v12.3 to 91.2% in v13.1. We applied strict criteria for considering a classifier prediction as consistent with the diagnosis. Without comprehensive molecular data in all cases, however, discrepant predictions cannot be definitively classified as incorrect.

In summary, we present a robust, automated classifier leveraging DNA methylation profiling for precise sarcoma diagnosis. The sarcoma classifier v13.1 is accessible at https://app.epignostix.com

***Supplementary figure 1***

**a** Copy number profiles in two distinct groups of angiosarcoma (AS) and difference in *MYC* amplification. Myc amplification occurs in mc AS_A while absent in mc AS_B. Chromosome 6q losses and 7 gains are more frequent in mc AS_B.

**b** Copy number profiles in two distinct groups of gastrointestinal tumors and differences in MGMT methylation and sex distribution. Mc GIST_AD exhibits a high frequency of 1p and 14q losses nor seen in mc GIST_PED. Mc GIST_PED occurs predominantly in female patients and tumors exhibit mostly methylation of MGMT.

***Supplementary figure 2***

Summary CNV profiles for 93 methylation classes covered in sarcoma classifier v13.1. X-axis gives chromosomal localization, y-axis provides percentage of tumors affected in the respective methylation class.

***Supplementary figure 3***

Calculated tumor cell content and predictions with a calibrated score >0.9 versus no prediction in 1547 cases of the four validation sets combined.

***Supplementary table 1***

Composition of the v13.1 reference set and comparison with the v12.3 reference set. The table provides abbreviations and full names of the tumor types included in v13.1 (n=4377) and v12.3 (n=1077). Tumors grouped to a single mc are indicated by boxes. * normal non-tumorous control tissues; ** one IFS_B included with IFS in v12.3; *** one LMS_UT included with LMO in v12.3; **** incorrectly termed CHORD_DD in v12.3; ***** removed from Sarcoma reference set because represented in brain tumor classifier 12.5 upward.

***Supplementary table 2***

Description of the methylation classes in v13.1. Amplifications and homozygous deletions observed in 10% or more of the cases within a mc are listed. Only extensive CNV encompassing at least half of the respective chromosomal arm are scored resulting in underscoring of circumscribed alterations which can be estimated in supplementary figure 1. Percentages given for CNV alterations, gene amplifications and homozygous deletions base on analyses of the original idat pairs resulting supplementary tables 3 to 6. All CNV observed in 60% or more of the respective groups are listed. In cases with fewer alterations the most frequently recurring lesion is shown if seen in 30% or more of the cases.

***Supplementary table 3***

1547 cases from four validation sets. Information included is original diagnosis, validation set, predictions of the 12.3 and 13.1 classifiers with calibrated scores and information on whether cases were eligible for the classifiers and whether a prediction was reached and whether the prediction matched with the original diagnosis.

## Equal Contribution

Natalie Jäger and David E. Reuss contributed equally to this work.

## Support

Supported by an Illumina research grant. The MASTER program is supported by the NCT Overarching Clinical Translational Trial Program, the NCT Heidelberg Molecular Precision Oncology Program, and DKTK.

## Data sharing statement

The training data (n=4377) set has been deposited at GHGA under reference number GHGAS64406795843199 and is available for download (raw idat files) upon signing a data transfer agreement.

## Acknowledgements

The authors thank Ulrike Lass, Joshua Trautner, Antje Habel and the DKFZ methylation core facility for skillful technical assistance. Supported by an Illumina research grant.

INFORM: We would like to express our sincere thanks to Carsten Maus, Erjia Wang (Genomics and Proteomics Core Facility, DKFZ). Lena Weiser, Gregor Warsow (Omics IT and Data Management Core Facility, DKFZ) for their highly dedicated support in data management and processing. Robert Autry, Gnanaprakash Balasubramanian, Christopher Previti and Rolf Kabbe (Division of Pediatric Neurooncology, DKFZ) for their sincere and dedicated contribution to the bioinformatics analyses. The INFORM program is financially supported by the German Cancer Research Center (DKFZ), several German health insurance companies, the German Cancer Consortium (DKTK), the German Federal Ministry of Education and Research (BMBF), the German Federal Ministry of Health (BMG), the Ministry of Science, Research and the Arts of the State of Baden-Württemberg (MWK BW); the German Cancer Aid (DKH), the German Childhood Cancer Foundation (DKS), RTL television, the aid organization BILD hilft e.V. (Ein Herz für Kinder) and the generous private donation of the Scheu family. We thank the NCT Sample Processing Laboratory and the DKFZ Next Generation Sequencing, Microarray, and Omics IT and Data Management Core Facilities for technical support.

